# A One-Arm Pilot Trial of a Telehealth CBT-Based Group Intervention Targeting Transdiagnostic Risk for Emotional Distress

**DOI:** 10.1101/2024.04.23.24306218

**Authors:** Sierra Flynt, Brandon Koscinski, Catherine Accorso, Ashley Knapp, Stephanie Gorka, Julie Suhr, Megan Austin, Nicholas P. Allan

## Abstract

The COVID-19 pandemic had a significant impact on mental health, straining an already overburdened healthcare system. A modular, transdiagnostic approach to treating psychopathology may be ideal to target common transdiagnostic risk factors for emotional distress and related disorders likely to be impacted by circumstances related to this once-in-a-lifetime environmental stressor. Anxiety sensitivity (AS), or fear of anxious arousal, intolerance of uncertainty (IU), or distress when confronted with uncertainty, and loneliness are three transdiagnostic risk factors impacted by the pandemic and robust predictors of emotional distress beyond that. We completed a pilot feasibility, acceptability, and utility trial of Coping Crew, our group, telehealth-delivered transdiagnostic treatment protocol in 17 participants who completed the intervention (*M* age = 22.00, *SD* = 4.46; 71% female). The intervention and study protocol were feasible to deliver and were rated as acceptable and useful to address intervention targets. Evidence was mixed regarding feasibility, acceptability, and usefulness of the mobile app component. Sixteen of 17 participants (94%) completed at least one survey a day on 80% of the days but only 6 participants (35%) completed at least 80% of the mobile app surveys delivered over the course of the intervention. Most participants rated use of the app as acceptable and relevant to psychological improvements made due to the intervention. Sizeable effect size reductions in transdiagnostic risk factors were found at post-intervention and maintained at 1- and 3-month follow-up, supporting next steps in the development of this modular transdiagnostic treatment.

---

The COVID-19 pandemic has had catastrophic impacts on not only physical health but also on mental health. In 2021, approximately two-thirds of individuals in the United States were experiencing moderate or severe psychological distress, as well as elevated anxiety (Cordaro et al., 2021; Khoury et al., 2021), depression (Bueno-Notivol et al., 2021), and loneliness (Moscona et al., 2021). The percentage of people who reported their anxiety as ‘high’ or ‘extremely high’ has quadrupled from 5% to 20% since the pandemic began and self-reported prevalence of depression has doubled (Dozois, 2021). Further, as the pandemic recedes, it is expected that people continue to seek mental health services at a higher rate than before the pandemic (Cromarty et al., 2020; Dozois, 2021). There is therefore a need for brief, modular telehealth interventions targeting emotional distress.

## Transdiagnostic Risk Factors

Disorder-specific cognitive-behavioral therapy (CBT) is widely used to treat emotional distress disorders despite how rare it is that people meet diagnostic criteria for only one disorder. In a national survey of 43,093 adults receiving treatment for mental health conditions in the United States, researchers found that 80.3% received two or more diagnoses (Kessler et al., 2005). Treatments could be more efficient and effective if focused on the core processes underlying multiple comorbid conditions (Hayes & Hofmann, 2017). Transdiagnostic risk factors are those that have been implicated in the etiology of multiple disorders (Struijs et al., 2021). These risk factors can directly influence the expression of symptoms in mental health disorders, affecting both their intensity and their change over time. Further, a growing body of literature demonstrates that transdiagnostic risk factors can be reduced through CBT and that reductions in these risk factors leads to improvement in emotional distress disorder symptoms (Roberge et al., 2022; Schaeuffele et al., 2022; Schmidt et al., 2014, 2016). Consequently, using CBT to target transdiagnostic risk factors may have broad effects across multiple disorders.

### Anxiety Sensitivity

Anxiety sensitivity (AS) is the fear of bodily sensations associated with anxiety (Reiss et al., 1986; Taylor & Cox, 1998). Lower-order dimensions of AS capture fears centered on physical, cognitive, and observable anxiety sensations and experiences. Elevated AS has been implicated as a risk factor for a wide range of maladaptive psychopathology, including anxiety disorders, depression, substance use and misuse, and suicidal urges (Allan et al., 2015; Lejuez et al., 2006; Olatunji & Wolitzky-Taylor, 2009; Stanley et al., 2018). Further, AS is positively associated with COVID-19-related worry, anxiety, functional impairment, and symptom severity (Rogers et al., 2021) and predicts fear of COVID-19 (Çelik et al., 2022) suggesting this will be an important elevated risk factor to target. Already, there is ample evidence that a brief AS intervention can reduce AS and associated psychopathology, including anxiety, depression, suicidal ideation, insomnia, and PTSD symptoms among others (Allan et al., 2015; Schmidt et al., 2014, 2017; Short et al., 2015, 2017).

### Intolerance of Uncertainty

Intolerance of uncertainty (IU) reflects “a dispositional characteristic that reflects an individual’s tendency to react negatively to uncertain situations, events, or outcomes, as well as the corresponding desire for certainty and predictability” (Carleton et al., 2012). There is strong evidence of a higher-order IU factor and mixed evidence for two lower-order IU factors, prospective IU or discomfort when confronted with future uncertainty, and inhibitory IU or difficulty tolerating uncertainty in the moment (Hale et al., 2016; Huntley et al., 2020). IU is implicated in the onset of various psychiatric diagnoses, including generalized anxiety disorder (GAD; Ren et al., 2021)and obsessive-compulsive disorder (OCD; Lind & Boschen, 2009). Changes in IU are related to increases in social anxiety, worry, and negative affect (M. Shapiro et al., 2020), along with changes in anxiety and depression symptoms across diagnostic categories (Boswell et al., 2013). Elevated IU during the COVID-19 pandemic has been identified as a risk factor for depression (Hamama-Raz et al., 2021; Moscona et al., 2021; Voitsidis et al., 2021), fear and anxiety (Dai et al., 2021), and suicidal ideation (Allan et al., 2021). IU is also positively associated with fear of COVID-19 (Çelik et al., 2022), COVID-19 worry, sensitivity to COVID-19-related symptoms, and COVID-19-related interference in daily activities due to worry (Allan et al., 2021). Already, there is emerging evidence that IU can be targeted in brief (M. O. Shapiro et al., 2023) and more intensive interventions (Dugas et al., 2022; van der Heiden et al., 2012).

### Loneliness

Loneliness is the subjective evaluation of one’s social relations as being inadequate or otherwise not fulfilling one’s social needs and desires (Calati et al., 2019; Hawkley & Cacioppo, 2010; Solmi et al., 2020). Loneliness is conceptualized as a unidimensional construct (Allen & Oshagan, 1995) and has been linked to various psychiatric diagnoses and related behavioral risk factors including anxiety disorders, depression (Moscona et al., 2021), and suicide attempts (McClelland et al., 2020). In a recent meta-analysis, loneliness was also associated with paranoia, and psychosis, as well as smoking, excess alcohol consumption, overeating, food restriction, low levels of physical activity, elevated mental health symptom severity, poor recovery prognosis, poor interpersonal functioning, and increased mortality (Solmi et al., 2020). Loneliness has also been identified as a potential risk factor for depression symptom severity, psychological distress, and financial worries during COVID-19 (Moscona et al., 2021). Almost half (41%) of a Canadian sample indicated that social isolation has had a significant negative impact on their mental health (Dozois, 2021). Brief interventions targeting perceived burdensomeness and thwarted belongingness, two constructs reflecting biases in perceived social relations have provided preliminary evidence that loneliness could be reduced through a brief intervention (Allan et al., 2018; Schmidt et al., 2023). Further, more intensive interventions have also proven successful in reducing loneliness (Käll et al., 2021).

## Development of a Modular CBT for Transdiagnostic Risk Factors Exacerbated by the COVID-19 Pandemic

Modular CBT treatments focus on using CBT principles to target an individual process or set of related processes underlying psychopathology (Schaeuffele et al., 2021). In this framework, risk processes—such as AS, IU, and loneliness—can be ideographically assigned to a client depending on their specific needs (Ellard et al., 2023). Modular CBT treatments, with their flexible and adaptable nature, provide an ideal framework for designing interventions that effectively target and address the transdiagnostic risk factors implicated in various emotional distress disorders (McHugh et al., 2009). The precision and adaptability of this approach align well with the overarching goal of precision medicine, offering personalized treatments that take into account individual variability in genes, environment, and lifestyle.

A central premise of CBT is that treatment gains are consolidated through out-of-session skills practice by the client (Kazantzis & Miller, 2022; Mausbach et al., 2010). However, unlike in-session activities, these activities have traditionally occurred without direct support which can reduce engagement in and gains from between-session activities (Kazantzis & Miller, 2022). The ubiquitous nature of smartphones and the general comfort with mobile apps provides an excellent platform to increase between-session client engagement. Generalization of the skills learned during CBT requires repeated exposure to brief treatment elements near the times they are needed. Ecological momentary assessment (EMA), delivered via mobile devices, offers an opportunity to easily implement skills practice as-needed by providing a monitoring system that can be used to determine when skills practice might be most beneficial. Ecological momentary interventions (EMIs) are built on top of EMA on the premise that ideal learning will occur by precisely providing the type of support needed in the time and place needed (Heron et al., 2017). Use of a mobile app also allows for accountability of completing behavioral exercises, another central component of most CBT-based treatments. In addition to improving the intervention, EMA also reduces retrospective recall bias, increases ecological validity, and allows for examination of short-term temporal dynamics (Shiffman et al., 2008).

## Current Study

Brief modular transdiagnostic interventions based in empirically supported CBT principles, delivered remotely, and supplemented with a mobile app offer an opportunity to efficiently and effectively target emotional distress common to a host of emotional distress disorders and exacerbated by the COVID-19 pandemic. AS, IU, and loneliness are three relevant risk factors to target given their associations with a wide range of psychopathology and distress due to the COVID-19 pandemic as well as evidence that they can be targeted through brief modules. We developed a five-session group intervention, Coping Crew, using a modular framework and guided by CBT principles. A mobile application was used to track mood, provide brief cognitive nudges, and monitor interoceptive exposure (IE) and behavioral experiments (BEs) done as homework. This study was designed to examine the acceptability, feasibility, and utility of Coping Crew and the associated mobile app as well as to provide preliminary effect size estimates and associations between transdiagnostic risk factors and emotional distress disorder symptom change over time.

## Methods

### Participants

Participants (*N*=17) included sixteen college students (94%) and one community member (6%) from a rural community in Southeast Ohio. Regarding self-reported race, 10 (59%) reported White, another two reported Asian and White (11.8%), another one reported Black (5.9%), and one reported Other (5.9%). All participants were non-Hispanic. Participants were 22.00 years old on average (standard deviation [*SD*] = 4.46) and 71% of participants were female.

### Procedures

This study was supported by an internal grant provided to the last author and registered on clinicaltrials.gov (CT# NCT05019053). Participants were recruited through email, social media, and digital flyers to clients of the university psychology clinic. To be eligible, participants had to a) report either elevated AS (Anxiety Sensitivity Index-3 [ASI-3] ≥ 24), IU (Intolerance of Uncertainty Scale-12 [IUS-12] ≥ 35), or loneliness (NIH Toolbox Loneliness scale ≥ 13), b) own a smartphone, and c) have internet access. Exclusion criteria included participation in other current lab interventions, reporting psychotic features, or imminent risk for suicide. See **Figure 1** for the Consort diagram reflecting study recruitment and retention. One hundred and seventy-six people completed the screening and 158 were eligible to participate. We contacted 96 participants sequentially from this list until we reached a sample of 20 participants. Three participants withdrew after consenting and 17 received Coping Crew. There was no single reason why participants withdrew: one participant did not want to download the mobile app, another participant was concerned that completing a standard I-9 form could lead to problems with their VISA, and another did not reside in Ohio and had to be withdrawn.

**Figure 1.**
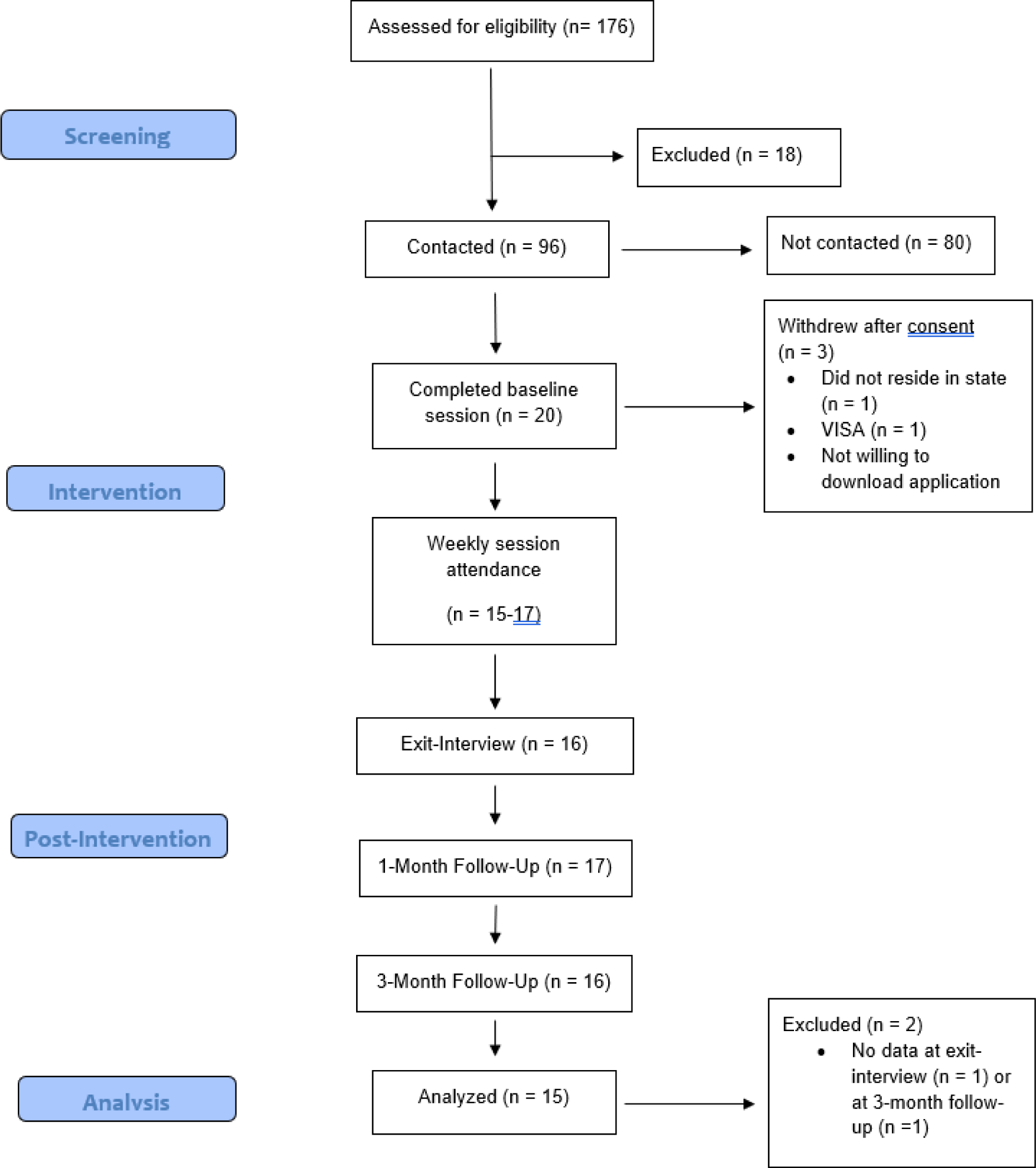
Consort diagram for Coping Crew.

Potential participants first completed a screening survey online using Qualtrics. Eligible and interested participants completed a baseline virtual appointment consisting of a structured diagnostic interview and a battery of self-report measures. Following the baseline appointment, participants received Coping Crew, consisting of five intervention sessions over six weeks. Next participants completed a battery of self-report measures at post-intervention and one- and three-month follow-up. At post-intervention, participants completed a semi-structured interview containing questions designed to capture perceived acceptability, usefulness, and effectiveness of the intervention, in addition to the user experience of the accompanying mobile application. Participants were compensated up to $130 for their participation.

### Coping Crew Intervention

Coping Crew was iteratively designed through focus groups and case series as a modular CBT intervention. As a starting point, content from Computerized Anxiety Sensitivity Treatment (Schmidt et al., 2014, 2017, 2023)(Schmidt et al., 2014, 2017, 2023) for the AS module and content from a recent psychoeducation-focused intervention for the IU module (M. O. Shapiro et al., 2023) was reviewed and adapted as necessary. Principles from Cully and Teten’s “A Therapist’s Guide to Brief Cognitive Behavioral Therapy” (Cully & Teten, 2008) were employed when shaping modules and homework.

The intervention consisted of five weekly 60-minute sessions delivered in a group format virtually via Microsoft Teams by two masters-level clinicians with a background in CBT. The first four sessions occurred weekly and there was a two week break between the fourth and fifth session to provide additional time for skills building. Four groups of the Coping Crew intervention, three groups of four participants and one group of six participants, were run from September 2021 to June 2022 with sessions at the same time each week there was a session to maintain continuity. Participants unable to attend a group session completed a make-up session with a clinician one-on-one prior to the next session.

### Coping Crew

Coping Crew is a modular CBT intervention that addresses emotional distress through a series of five remote group sessions. The program uses the modular CBT approach by focusing on a different topic each week: AS, IU, and Loneliness. The first session introduces the program, establishes group norms, and promotes voluntary intra-group sharing to enhance corrective feedback. The CBT approach is explained, and participants practice relaxation exercises.

The next three sessions each target a transdiagnostic risk factor for emotional distress using a similar format. In all sessions, homework from the preceding week is reviewed in session and space is given to address participant questions or concerns. Session two next introduces anxiety, its role as the body’s alarm system, and how to challenge misperceptions about anxiety sensations. Behavioral experiments, such as rapid breathing to mimic shortness of breath or running in place to mimic a rapidly beating heart are assigned to reinforce that anxiety sensations are benign. Session three introduces IU, its role in causing and maintaining anxiety, and common misperceptions about uncertainty. Participants are urged to challenge their own beliefs about uncertainty. Finally, participants design behavioral experiments that encourage confronting uncertainty. For example, participants chose to delegate a task to a co-worker, answer a question in class even if they did not know the answer with complete certainty. In session four, the focus shifts to social isolation experienced due to the pandemic and any accompanying loneliness. Strategies based on Behavioral Activation (BA) are introduced to encourage positive social interactions and challenge beliefs that inhibit interpersonal connection. Homework includes choosing social activities such as making small talk with a stranger or reaching out to an old friend.

The final session wraps up the intervention by revisiting all topics covered. In this session, the information covered is summarized, self-reflection is encouraged, strategies to maintain treatment gains are discussed, and goal setting and values are introduced. Participants are provided with resources related to further obtaining mental health care if desired as well as resources to utilize in the event of a mental health crisis. This modular CBT-based intervention is designed to promote resilience in the face of anxiety, accompanying anxiety sensations, and social isolation.

### Mobile Intervention Component

The mobile component of the intervention was delivered using a HIPAA-secured mobile application available from Metricwire (metricwire.com) named mEMA. Over a 12-hour window, a static survey was delivered and then three additional surveys were quasi-randomly delivered no closer than an hour and a half apart over the remaining time. Participants completed a set of self-report questionnaires in the morning and visual analog scales (VAS) across all assessments. Cognitive challenges were delivered if VAS ratings were ≥ 5 on a 10-point scale. These cognitive challenges comprised “key takeaways” from each week’s intervention topic developed by members of the group, beginning in week 2. These messages were challenges to maladaptive cognitions related to each session’s content. For example, AS-specific challenges included “anxiety is just a feeling” and “the anxiety I feel now will pass.” IU-specific challenges included “even if things go off plan, I can adjust for that” and “uncertainty doesn’t mean something bad is going to happen” and loneliness-specific challenges included “avoiding social interactions won’t improve feeling alone.” After week 2, whenever a participant reported anxiety or stress ≥ 5, one of these key takeaways was delivered randomly from a que of takeaways generated in-session. AS and IU-related key takeaways were randomly selected from after week 3 with a 75% likelihood of drawing an IU-related key takeaway. After week 4, AS and IU key takeaways were equally delivered whenever a participant reported elevated anxiety or stress and loneliness key takeaways were delivered whenever a participant reported elevated depression. The prompts for each emotional distress and the corresponding behavioral experiments evolved with the progression of the intervention, giving participants an interactive, personalized, and comprehensive experience.

### Feasibility, Acceptability, and Utility Measures

The study employs a custom set of evaluative measures to gauge the feasibility of the intervention and associated assessment battery and acceptability and utility of the intervention and mobile app. **Feasibility** was determined by attendance rates for assessment and intervention sessions and rates of survey completion via mobile app. **Acceptability** was measured by Perceived **Acceptability of the Intervention** and **Acceptability of the Mobile Application** which participants’ attitudes towards the intervention’s impact on AS, IU, loneliness, anxiety, stress, and depression, as well as the anticipated sustainability of improvements. Perceived **Utility of the Services Provided** included questions focusing the impact of the intervention on reducing anxiety, stress, and loneliness and using the mobile app to track weekly activities. All items were assessed on a 5-point Likert-like scale with higher scores indicating greater agreement with the item.

### Assessment Measures

**Anxiety Sensitivity Index-3.** The ASI-3 (Taylor et al., 2014) is an 18-item measure that assesses fear of physical, cognitive, and observable anxiety sensations by measuring three AS subfactors (physical, cognitive, and social concerns). Items are assessed on a 5-point Likert-like scale with higher scores indicating greater AS. In the current study, reliability for the ASI-3 from baseline to follow-up was excellent (Cronbach’s alpha [α] = .91-.95).

### Intolerance of Uncertainty Scale Short Form

The IUS-12 is a 12-item measure that assesses the degree to which individuals can tolerate the uncertainty of ambiguous situations, cognitive and behavioral responses to uncertainty, perceived implications of uncertainty, and attempts to control the future through two factors: prospective anxiety and inhibitory anxiety (Carleton et al., 2007). Items are assessed on a 5-point Likert-like scale where 1 indicates that the respondent does not find the statement to be characteristic of themselves and 5 indicates that the respondent finds the statement to be very characteristic of themselves. In the current study, reliability for the IUS-12 from baseline to follow-up was good to excellent (Cronbach’s alpha ranged from.83 at week 1 to 93 at baseline).

### National Institute of Health Toolbox: Adult Social Relationship Scales: Loneliness

The NIH Toolbox Loneliness Scale (Cyranowski et al., 2013) is 5-item measure within the NIH Toolbox Adult Social Relationship Scales that assesses loneliness, conceptualized as the perception that one is lonely or socially isolated from others. Items are assessed on a 5-point scale with 1 indicating the respondent never has the experience and 5 indicating that the respondent always has the experience. In the current study, reliability for the NIH Loneliness scale from baseline to follow-up was good to excellent (Cronbach’s alpha ranged from .87 at week 2 to.96 at baseline).

### PROMIS Negative Affect Scales: Anxiety & Depression

The Patient Reported Outcomes Measurement Information System (PROMIS) Negative Affect scales include measures for assessing anxiety and depression (Pilkonis et al., 2011). The PROMIS Anxiety item bank consists of 29 items and the PROMIS Depression item bank consists of 28 items. We utilized the short forms of both measures, containing 8 items each. Reliability for the PROMIS Depression scale from baseline to follow-up was excellent (Cronbach’s alpha ranged from .95 at week 1 and post-intervention to.97 at 3-month follow-up).

### COVID Impact Battery: COVID-19 Worry Scale

The COVID Impact Battery (CIB) captures the influence of the COVID-19 pandemic on individual mental health across three COVID-19-related scales: behavior, worry, and disability (Schmidt et al., 2022). The COVID-19 Worry scale is an 11-item measure that assesses COVID-related stress, associated with avoidance of infection via safety behaviors, self-checking, and reassurance-seeking (i.e. utilization of the healthcare system), as well as worry about socioeconomic impacts of the pandemic (Taylor et al., 2020). Items are assessed on a 0 to 4 scales with higher scores capturing increased worry. In the current study, reliability for the CIB Worry scale from baseline to follow-up was good to excellent (Cronbach’s alpha ranged from .84 at baseline to.92 at 3-month follow-up).

### EMA Measures

During the intervention, participants completed daily surveys over the course of six weeks. Each day participants received four brief surveys. The first survey was sent at the beginning of a 12-hour period predetermined by the client and clinician, followed by three surveys delivered randomly throughout the remaining 12-hour window. All surveys were delivered at least 90 minutes apart. The purpose of the personalized time period was to maximize participant engagement, by increasing likelihood of survey completion via convenience. The brief surveys were designed to take approximately one minute to complete. EMA measures included the CIB and four Visual Analog Scales (VAS): Anxiety, Depression, Stress, and Loneliness. VAS measures were formatted as slider bars for participants to rate their levels on a 1 to 100 scale (e.g., How anxious are you now?) with higher scores indicating more severe levels of the rated construct.

### Data Analytic Plan

This study was a one-arm pilot acceptability and feasibility study, registered on clinicaltrials.gov (NCT#05019053). Reporting was done in accordance with the Consolidated Standards of Reporting Trials (CONSORT) guidelines, adopted for one-arm trials (Eldridge et al., 2016). Acceptability, feasibility, and utility ratings were first calculated by examining mean item-level scores across several measures modified to capture participant ratings of acceptability, feasibility, and utility. Acceptability and feasibility were also assessed through monitoring recruitment, retention, and use of the mobile app. Following this, repeated measures ANOVAs were conducted to examine changes in mean AS, IU, anxiety, depression, and worry related to the COVID-19 pandemic from baseline to post-intervention and 1-month follow-up. Mauchly’s test was used to determine if the assumption of sphericity was met. If it was not met, the Greenhouse Geisser statistic was used to determine the significance of the tests of within-subjects effects. Because this trial was not designed to be powered to find all but the largest of effect sizes, we were primarily interested in standardized effect size estimates (*d*w) which was calculated as the change in scores divided by the pooled baseline standard deviation.

## Results

### Eligibility Criteria Met for Enrollment

Eligibility criteria included elevations of at least 1 *SD* above the mean on at least one risk factor: AS, IU, or loneliness. At baseline, thirteen participants had elevated ASI-3 scores. Seventeen participants had elevated IUS-12 scores. Finally, six participants had elevated NIH Loneliness scores.

### Acceptability, Feasibility, and Utility

Acceptability, feasibility, and utility were examined across attendance and performance metrics as well as response to Likert-like questions. Twenty participants were consented and completed the baseline session; however, three participants withdrew and 17 participants were allocated to receive Coping Crew (85%). All 17 participants attended one or more treatment sessions and 15 attended all five intervention sessions. All 17 participants completed their baseline and 1-month follow-up survey batteries. Only one participant did not complete the 3-month follow-up battery. Two participants missed session two, one participant missed session three, and one participant missed session four. No participant missed more than a single session. This suggests delivery of the intervention and corresponding assessment protocol are feasible. Performance on the daily surveys via the mobile app was mixed. Only 6 (35%) of 17 participants completed 80% or more of the surveys delivered to their mobile app. However, 16 of 17 (94%) completed at least one survey daily 80% of the days. Participants completed an average of 13 (2 to 23 were completed) behavioral experiments which had been assigned as daily homework.

Participants rated perceived efficacy of the intervention component delivered by clinicians across 10 items with 1 indicating they strongly disagreed with the item and 5 indicating they strongly agreed (see Top Panel of Table 1). The average item rating was 4.23 (3.75-4.63), reflecting that participants agreed, on average that the intervention was acceptable in a variety of ways, including for anxiety, stress, and depression. Of note, participants strongly agreed (*M* = 4.63, *SD* = .62) that they would recommend this approach to others who are lonely and anxious/stressed.

**Table 1.**
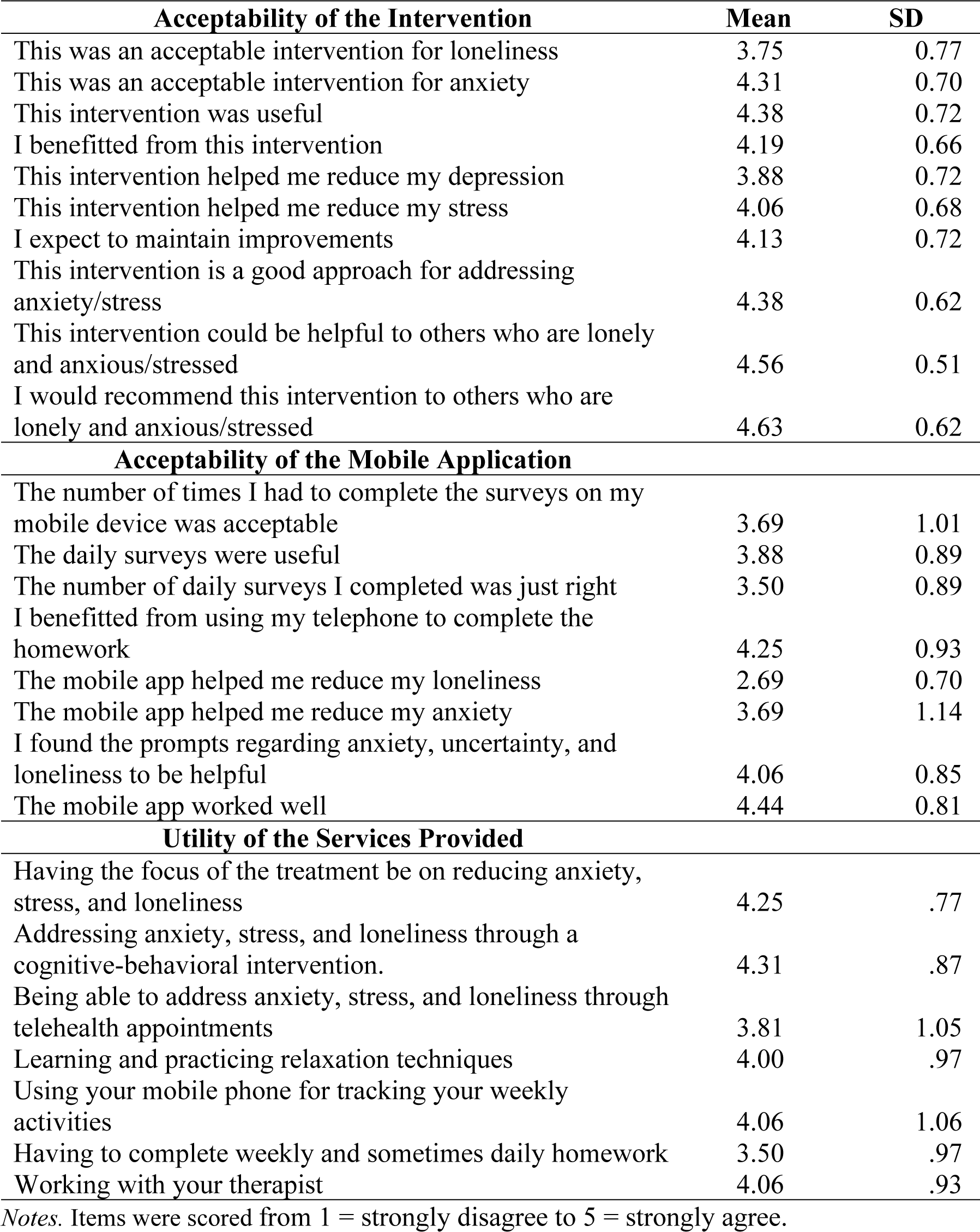
Ratings of Acceptability.

| **Acceptability of the Intervention** | **Mean** | **SD** |
| --- | --- | --- |
| This was an acceptable intervention for loneliness | 3.75 | 0.77 |
| This was an acceptable intervention for anxiety | 4.31 | 0.70 |
| This intervention was useful | 4.38 | 0.72 |
| I benefitted from this intervention | 4.19 | 0.66 |
| This intervention helped me reduce my depression | 3.88 | 0.72 |
| This intervention helped me reduce my stress | 4.06 | 0.68 |
| I expect to maintain improvements | 4.13 | 0.72 |
| This intervention is a good approach for addressing anxiety/stress | 4.38 | 0.62 |
| This intervention could be helpful to others who are lonely and anxious/stressed | 4.56 | 0.51 |
| I would recommend this intervention to others who are lonely and anxious/stressed | 4.63 | 0.62 |
| **Acceptability of the Mobile Application** |  |  |
| The number of times I had to complete the surveys on my mobile device was acceptable | 3.69 | 1.01 |
| The daily surveys were useful | 3.88 | 0.89 |
| The number of daily surveys I completed was just right | 3.50 | 0.89 |
| I benefitted from using my telephone to complete the homework | 4.25 | 0.93 |
| The mobile app helped me reduce my loneliness | 2.69 | 0.70 |
| The mobile app helped me reduce my anxiety | 3.69 | 1.14 |
| I found the prompts regarding anxiety, uncertainty, and loneliness to be helpful | 4.06 | 0.85 |
| The mobile app worked well | 4.44 | 0.81 |
| **Utility of the Services Provided** |  |  |
| Having the focus of the treatment be on reducing anxiety, stress, and loneliness | 4.25 | .77 |
| Addressing anxiety, stress, and loneliness through a cognitive-behavioral intervention. | 4.31 | .87 |
| Being able to address anxiety, stress, and loneliness through telehealth appointments | 3.81 | 1.05 |
| Learning and practicing relaxation techniques | 4.00 | .97 |
| Using your mobile phone for tracking your weekly activities | 4.06 | 1.06 |
| Having to complete weekly and sometimes daily homework | 3.50 | .97 |
| Working with your therapist | 4.06 | .93 |

*Notes.* Items were scored from 1 = strongly disagree to 5 = strongly agree.

Participants rated perceived efficacy of the mobile app components of the intervention across 8 items using the same 1-5 Likert scale (see Middle Panel of Table 1). The average rating was 3.77 (2.69-4.44), indicating participants agreed the mobile app was acceptable in a variety of ways, including finding the daily surveys useful (*M* = 3.88, *SD* = .89), and using their telephone to complete homework (*M* = 4.25, *SD* = .93). Participants strongly agreed (*M* = 4.44, *SD* = .81) that the mobile app worked well. It should be noted that the average level of agreement on whether the mobile app was helpful for loneliness was neutral, meaning participants neither agreed or disagreed that it was helpful.

### Preliminary Data Analysis

We examined differences in baseline mean scores between participants who were missing data versus participants who were not missing any data. A significant difference in PROMIS Depression scale scores was found *t*(15) = -2.06, *p* = .03. Participants who provided data at all assessment periods had lower depression scores (*M* = 15.50, *SD* = 5.66; *n* = 8) than did participants who did not provide data at all assessment periods (*M* = 21.56, *SD* = 6.39; n = 9). No other differences were found.

### Baseline Descriptive Statistics and Correlations

Table 1 contains correlations between baseline variables as well as sample means. ASI-3, IUS-12, and NIH Loneliness scores were significantly correlated (*r*s = .51-.72, *p*s < .05). All three risk factors were associated with the PROMIS Anxiety scale (*r*s = .55-.79, *p*s < .05). The NIH Loneliness scale was also associated with scores on the PROMIS Depression scale (*r* = .64, *p* < .05) and COVID-19 Worry scale (*r* = .50, *p* < .05).

### Repeated Measures ANOVAs

See Table 3 for repeated measures ANOVAs examining differences in mean scores across baseline, post-intervention, and follow-up assessment sessions.

**Table 2.** Bivariate correlations between outcome measures at baseline.

| **Measure** | **1** | **2** | **3** | **4** | **5** | **6** | **7** | **8** |
| --- | --- | --- | --- | --- | --- | --- | --- | --- |
| 1. ASI-3 | - | .72** | .63** | .69** | .44 | .42 | -.09 | -.11 |
| 2. IUS-12 | - | - | .51* | .55* | .19 | .17 | -.09 | .197 |
| 3. NIH Loneliness | - | - | - | .68* | .64** | .50* | .10 | -.02 |
| 4. PROMIS Anxiety | - | - | - | - | .55* | .43 | -.12 | -.16 |
| 5. PROMIS Depression | - | - | - | - | - | .40 | .18 | -.07 |
| 6. CIB Worry | - | - | - | - | - | - | .16 | -.57* |
| 7. Age | - | - | - | - | - | - | - | .34 |
| 8. Sex | - | - | - | - | - | - | - | - |
| Mean | 34.12 | 53.88 | 14.94 | 25.29 | 18.71 | 16.29 | 22.35 | - |
| SD | 12.14 | 9.85 | 5.37 | 6.89 | 6.64 | 7.85 | 4.46 | - |

*Notes.* ASI-3 = Anxiety Sensitivity Index-3; IUS-12 = Intolerance of Uncertainty Scale-12; NIH = National Institutes of Health; PROMIS = Patient-Reported Outcomes Measurement Information System. CIB = Coronavirus Information Battery
\* $p \leq .05$
\*\* $p \leq .01$

**Table 3.** Repeated measures ANOVA for study outcomes.

| **Variables** | Baseline (1) |  | Post-Intervention (2) |  | Month 1 (3) |  | Month 3 (4) |  | Overall F |
| --- | --- | --- | --- | --- | --- | --- | --- | --- | --- |
|  | **M** | **SD** | **M** | **SD** | **M** | **SD** | **M** | **SD** |  |
| ASI-3 | 34.12<sup>2,4</sup> | 14.14 | 17.38<sup>1</sup> | 14.15 | 21.53 | 14.16 | 20.06<sup>1</sup> | 13.91 | 9.95* |
| IUS-12 | 53.88<sup>2,3,4</sup> | 9.85 | 30.69<sup>1</sup> | 9.51 | 31.41<sup>1</sup> | 9.47 | 53.88<sup>1</sup> | 9.85 | 51.06* |
| NIH Loneliness | 14.94 | 5.37 | 11.00 | 4.55 | 11.81 | 4.40 | 12.71 | 5.18 | 3.53* |
| PROMIS Anxiety | 25.29 | 6.89 | 21.56 | 7.13 | 21.59 | 6.53 | 23.00 | 6.39 | 1.43 |
| PROMIS Depression | 18.71 | 6.64 | 15.88 | 6.23 | 18.53 | 7.05 | 18.69 | 9.28 | .38 |
| CIB Worry | 16.29 | 7.85 | 12.56 | 9.19 | 13.35 | 8.92 | 13.12 | 9.77 | 1.45 |

*Notes.* ASI-3 = Anxiety sensitivity index-3; IUS-12 = Intolerance of uncertainty-12; CIB = Covid Impact Battery. <sup>1-4</sup>Reflect group
differences when comparing each group to the other groups.
\* $p < .05$ .

Repeated measures ANOVAs were conducted to examine differences in ASI-3 scores at baseline, 1-month, and 3-month follow-up. Mauchly’s test indicated that the assumption of sphericity was not met for the ASI-3; therefore the Greenhouse Geisser statistic was used to determine the significance of the tests of within-subjects effects. There was a statistically significant difference in ASI-3 mean scores across baseline, post-intervention, one month and three month follow-up (*F*(2.31) = 9.95, *p* ≤ .001. ASI-3 scores were significantly higher at baseline compared to post-intervention (*d* = 1.12), 1-month follow-up (*d* = .83), and 3-month follow-up (*d* = .94) scores.

Mauchly’s test indicated that the assumption of sphericity was not met for the IUS-12; therefore the Greenhouse Geisser statistic was used to determine the significance of the tests of within-subjects effects. There was a statistically significant difference in IUS-12 mean scores across baseline, post-intervention, one month and three month follow-up (*F*(2.194) = 51.06, *p* ≤ .001. IUS-12 scores were significantly higher at baseline compared to post-intervention (*d* = 2.5), 1-month follow-up (*d* = 2.17), and 3-month follow-up (*d* = 2.03).

Mauchly’s test indicated that the assumption of sphericity was not met for the NIH Loneliness Scale, x^2 (5) = 8.34, *p* = .14, therefore the Greenhouse Geisser statistic was used to determine the significance of the tests of within-subjects effects. There was a significant difference between NIH Loneliness mean scores across baseline, post-intervention, 1-month follow-up, and 3-month follow-up. (*F*(2.10) = 3.53, *p* = .04. There were no significant differences in loneliness scores across measurement occasions; however, the effect size reductions from baseline to post-intervention (*d* = .66), 1-month follow-up (*d* = .44), and 3-month follow-up (*d* = .51) were all medium in magnitude.

Mauchly’s test indicated that the assumption of sphericity was met for PROMIS Anxiety, x^2 (5) = 13.70, *p* = .02. therefore, the Mauchly’s W statistic was used to determine the significance of the tests of within-subjects effects. There were no significant differences between PROMIS Anxiety scores across timepoints (*F*(3) = 1.43, p = .25. Small to medium effect size reductions were found from baseline to post-intervention (*d* = .44), 1-month follow-up (*d* = .51), and 3-month follow-up (*d* = .32).

Mauchly’s test indicated that the assumption of sphericity was met for PROMIS Depression, x^2 (5) = 17.62, *p* = .004. therefore the Mauchly’s W statistic was used to determine the significance of the tests of within-subjects effects. There were no significant differences between PROMIS Depression scores across timepoints (*F*(3) = .38, *p* = 77. Further, effect size reductions were not meaningful: baseline to post-intervention d = .25, baseline to 1-month follow-up *d* = .01, baseline to 3-month follow-up *d* = -.01.

Mauchly’s test indicated that the assumption of sphericity was met for the CIB Worry Scale, x^2 (5) = 17.522, *p* = .004, therefore the Mauchly’s W (Sphericity assumed) statistic was used to determine the significance of the tests of within-subjects effects. There were no significant differences between PROMIS Depression scores across timepoints (*F*(3) = 1.45, *p* = .24. Despite this, small-to-medium effect size reductions were found from baseline to post-intervention (*d* = .44), 1-month follow-up (*d* = .36), and 3-month follow-up (*d* = .43).

## Discussion

This study was a preliminary feasibility, acceptability, and utility one armed pilot trial for Coping Crew. Coping Crew is a modular group CBT-based transdiagnostic intervention. To encourage use of in-session skills developed, we designed a mobile app to supplement the treatment. The intervention and assessment battery were feasible, acceptable, and had utility in addressing the targeted transdiagnostic risk factors. Although the mobile app was rated as acceptable and had utility in aiding delivery of the intervention, rates of completion were lower than we had proposed as our threshold for acceptability. Effect sizes capturing pre-to post-intervention change were promising for AS, IU, anxiety, and loneliness. Effect sizes capturing change for depression were not.

We determined that Coping Crew was acceptable and feasible based on recruitment and retention rates as well as ratings on measures assessing agreement that aspects of the intervention were acceptable and useful. We retained 85% of participants (17 of 20) consented to receive the intervention and 88% (15 of 17) of participants attended all five intervention sessions as well as the month 3 follow-up and 94% (16) attended the month 1 follow-up. On average, the intervention content was rated as acceptable. One item to highlight: on average, participants strongly agreed that they would recommend this intervention to others who are lonely and anxious/stressed. This suggests that client perceptions of Coping Crew align with the intended purpose of the treatment.

In contrast to the almost universally positive perceptions of the telehealth component, perceptions about the mobile app were more mixed. We selected, a priori, that if 80% of the participants completed at least 80% of the surveys delivered to their phone, we would consider that as evidence that the mobile app was a feasible accompaniment to the intervention. We only found six participants (35%) met this threshold. However, we did find that 94% of participants completed at least one survey on more than 80% of the days. Evidence for the acceptability and utility of the mobile app was mixed. On average, participants agreed they completed an acceptable amount of surveys daily, that they benefitted from using their phone to complete their homework and that the prompts they received regarding anxiety, uncertainty, and loneliness were helpful. We did learn from feedback to open- and close-ended questions that one participant was opposed to use of the mobile app. This participant disagreed to strongly disagreed that the mobile app and using the app to track their mood was a useful adjunct to the treatment. It is possible that there may be barriers to engaging with mobile apps for some participants. For example, the elderly, those in low-income groups, and rural populations might not have the necessary resources or digital literacy skills to benefit from an app (Choi et al., 2022). Others may be concerned about data privacy and confidentiality (Robillard et al., 2019). There is an emerging literature on engagement efforts for mHealth interventions; more is needed to understand what factors might suggest a research participant or clinical patient would not benefit from using a mobile app to track progress and whether there are acceptable substitutes.

Effect sizes obtained in one armed trials must be interpreted with caution due to the lack of a control condition. However, the lack of significant reductions in depression symptoms following Coping Crew suggested a need to consider whether Coping Crew should be pursued as a transdiagnostic treatment across emotional disorders. There is some evidence that this sample was not experiencing elevated loneliness. Only six participants endorsed elevated loneliness at baseline and ratings of acceptability for intervention components were generally lower in terms of how it helped participants with their loneliness. This includes our only average rating of neutral for “the mobile app helped me with my loneliness.”

Loneliness has repeatedly been identified as a potent risk factor for depression. Studies, such as those conducted by Hawkley et al. (2010) and Cacioppo et al. (2006), have firmly established that loneliness is associated with increased depressive symptomatology. Given this connection, the lack of changes in depression scores could be influenced by the initial sample characteristics. Specifically, given only six participants endorsed elevated loneliness at baseline, and loneliness is a stronger correlate of depression than AS and IU are, this might have constrained the potential to observe significant reductions in depression symptoms following the intervention.

Another possible explanation is that the intervention includes a wide enough array of transdiagnostic risk factors impacting anxiety (AS, IU) but not a wide enough array of risk factors for depression (loneliness). If this were the case, then consideration would need to be given as to whether to include modules to more fully address depression or whether to include less modules to focus only on anxiety. A Sequential Multiple Assignment Randomized Trials (SMART) may be an ideal approach to test whether individuals experiencing elevated loneliness at baseline would benefit from the loneliness module after the AS and IU modules. Participants could be randomized to conditions only in the event they elevate on the associated risk factor.

A further benefit of validating Coping Crew via a SMART trial is the ability to test the inherent modularity of the intervention. Modular treatments have emerged as a promising approach that bridges the gap between traditional psychotherapy practices, driven by clinician intuition, and manualized treatments, characterized by rigid protocols (Chorpita et al., 2005). Whereas traditional approaches relied on clinicians’ adaptability to meet clients’ evolving needs, manualized treatments provided structure at the cost of flexibility. In contrast, modular treatments allow for manualized treatments to be utilized based solely on participants current needs (Chorpita et al., 2005). Modular frameworks acknowledge the finite nature of therapeutic tools and offer the potential for “continuous scaling” from a common treatment framework (Chorpita et al., 2005). For instance, in the development of Coping Crew, an intervention currently under development, principles derived from both first and third-wave cognitive-behavioral therapy (CBT) as well as mindfulness practices are utilized to explore the intricate interplay among thoughts, feelings, and behaviors, with a specific focus on the psychological modifiability of thoughts and behaviors (Hofmann et al., 2012).

## Limitations

Despite grounding our study in prior literature, several limitations should be acknowledged. First, reliance on self-report measures introduces potential biases associated with memory processes and response tendencies. Whereas real-time data collection via EMA may mitigate some biases, further exploration of alternative measurement approaches is warranted. Second, the small sample size and convenience sampling resulting in a primarily white non-Hispanic and female sample limit the generalizability of our findings. Further, the lack of a control group limits interpretation of change. Multi-arm fully powered RCT designs are needed to fully explore efficacy of Coping Crew as well as identify mechanisms underlying treatment effects. These limitations provide directions for future research and further highlight the need for an RCT comparing Coping Crew to a well-validated control condition.

## Conclusion

This study served as a one-arm pilot feasibility, acceptability, and utility trial. Results were promising for the intervention and assessment protocol. Considerations for next steps include how to increase engagement in the mobile app and fitting intervention modules to sample characteristics. These findings leave us eager to conduct next stages in this modular transdiagnostic intervention that includes a mobile app component to supplement the intervention with limited clinical effort.

## Data Availability

Data are available through correspondence with Dr. Allan and with an appropriate data use agreement obtained.

